# Right bundle branch pacing: criteria, characteristics and outcomes

**DOI:** 10.1101/2022.11.16.22282347

**Authors:** Marek Jastrzębski, Grzegorz Kiełbasa, Paweł Moskal, Agnieszka Bednarek, Marek Rajzer, Karol Curila, Haran Burri, Pugazhendhi Vijayaraman

## Abstract

**Background:** Targets for right-sided conduction system pacing (CSP) include His bundle and right bundle branch. ECG patterns, diagnostic criteria and outcomes of right bundle branch pacing (RBBP) are not known.

**Objective:** Our aims were to delineate electrocardiographic and electrophysiological characteristics of RBBP and to compare outcomes between RBBP and His bundle pacing (HBP).

**Methods:** Patients with confirmed right CSP were divided according to the conduction system potential to QRS interval at the pacing lead implantation site. Six hypothesized RBBP criteria as well as pacing parameters, echocardiographic outcomes and all-cause mortality were analyzed.

**Results:** All analyzed criteria discriminated between HBP and LBBP: double QRS transition during threshold test, selective paced QRS different from conducted QRS, stimulus to selective QRS > potential-QRS, small increase in V_6_ RWPT during QRS transition, equal capture thresholds of CSP and myocardium, and stimulus-V_6_ R-wave peak time (V_6_ RWPT) > potential-V_6_ RWPT (adopted as diagnostic standard). Per this last criterion, RBBP was observed in 19.2% (64/326) patients who had been targeted for HBP, present mainly among patients with potential to QRS < 35 ms (90.6%, 48/53) and occasionally in the remaining patients (5.6%, 16/273). RBBP was characterized by longer QRS (by 10.5 ms), longer V_6_ RWPT (by 11.6 ms) and better sensing (by 2.6 mV) compared to HBP. During median follow-up of 29 months, no differences in capture threshold, echocardiographic outcomes or mortality were found.

**Conclusions:** RBBP is a distinct CSP modality that is frequently observed when the pacing lead is positioned more distally along the right conduction system.

## Introduction

Several studies have investigated characteristics and outcomes of the two major conduction system pacing CSP methods: His bundle pacing (HBP) and left bundle branch pacing (LBBP).^1-6^ However, right bundle branch (RBB) is also an accessible right-sided pacing target that might result in rapid engagement of the whole conduction system. No electrophysiological criteria for the diagnosis of RBB pacing (RBBP) have ever been proposed and no data regarding ECG characteristics, pacing parameters and impact on heart function and clinical outcomes are available. Moreover, prevalence of RBBP during attempted HBP is not known.

On the basis of our insights from parahisian pacing maneuvers performed during electrophysiological studies (**Figures 1 and 2**) and prior research regarding HBP and LBBP criteria,^7, 8^ we hypothesized that permanent RBBP may be characterized by several features that separate it from HBP.

**Figure 1.**
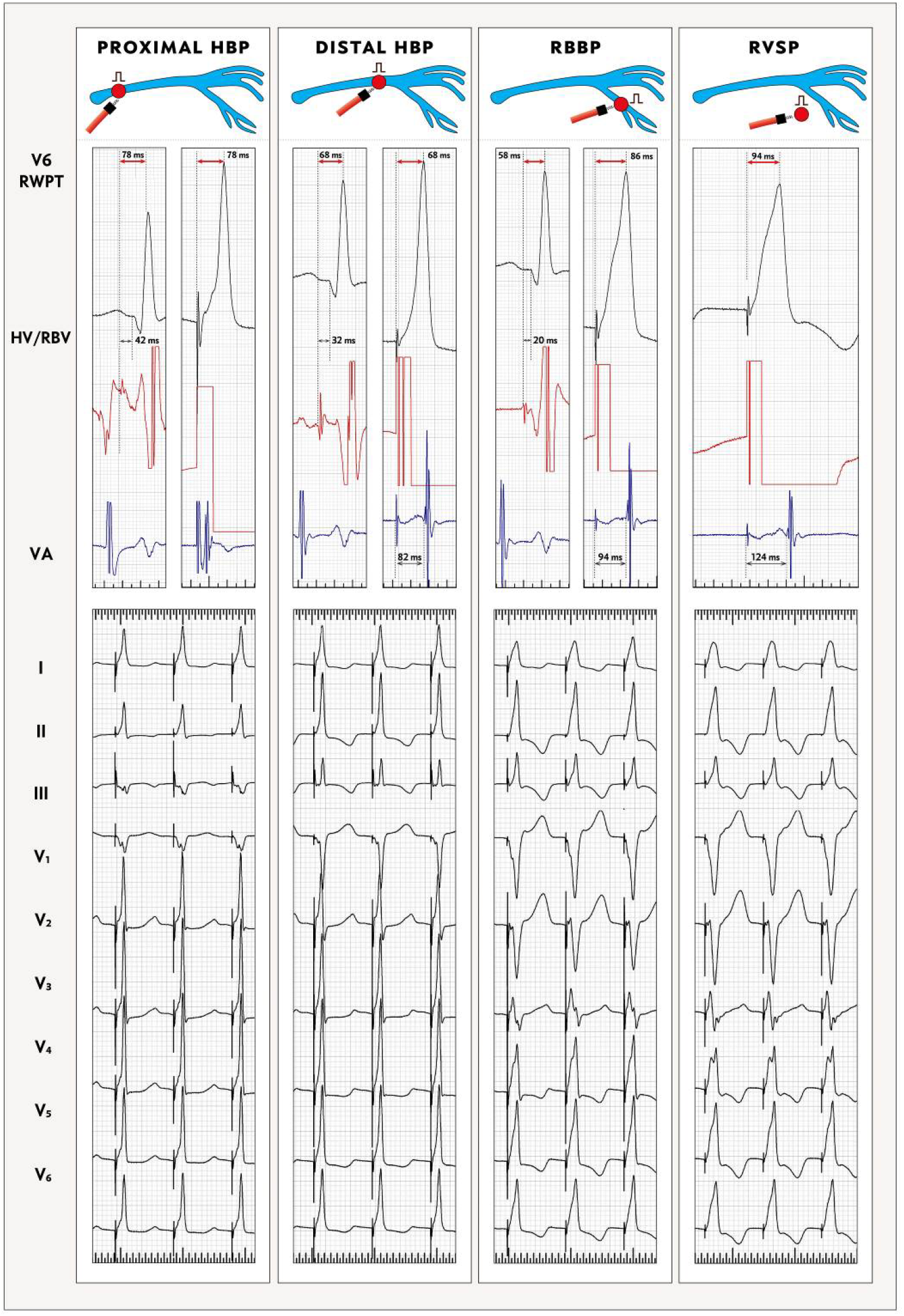
Pacing at different levels of the right-sided conduction system. The electrophysiology catheter was positioned in the His bundle area and the pacing site was moved from a proximal to a distal position. Proximal His bundle pacing (HBP), characterized by a large A wave, resulted in the paced V_6_ RWPT (interval from the stimulus to R-wave peak in lead V_6_) equal to the native V_6_ RWPT (measured from the HB potential). Distal HBP also resulted in equal native V_6_ RWPT and paced V_6_ RWPT. In contrast, right bundle branch pacing (RBBP), characterized by potential to QRS of 20 ms and native V_6_ RWPT of 58 ms, resulted in much longer paced V_6_ RWPT of 86 ms (for explanation see text). With right ventricular septal pacing (RVSP), the paced V_6_ RWPT was further increased to 94 ms and ventriculo-atrial interval (VA) substantially prolonged. As the pacing site becomes more distal, the paced QRS shows bigger impact of direct septal myocardial depolarization.

**Figure 2.**
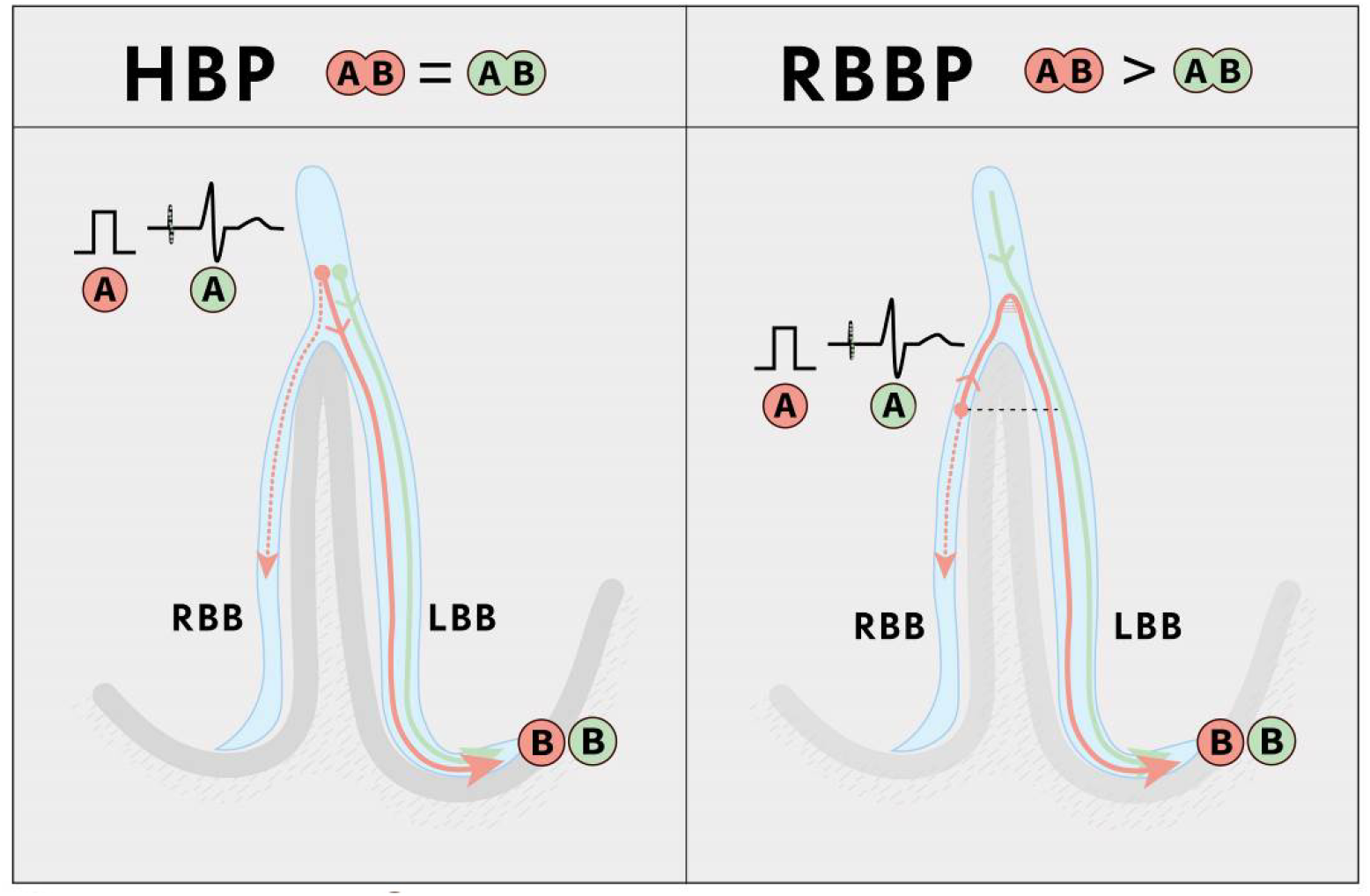
Difference in activation of the lateral wall of left ventricle (LV) during His bundle pacing (HBP) and right bundle branch pacing (RBBP). During HBP, the activation pathway from the point of HB capture / HB potential recording (A) to the activation of the lateral wall of the left ventricle (B), approximated by the V_6_ R-wave peak time (V_6_ RWPT) is the same during pacing and intrinsic conduction (paced V_6_ RWPT measured from stimulus = native V_6_ RWPT measured from HB potential). During native rhythm, the RBB potential to V_6_ RWPT is short (as V_6_ RWPT is determined by fast LBB conduction), while during RBBP, the activation pathway to the lateral wall of the left ventricle is long due to retrograde conduction via the RBB followed by anterograde conduction via the LBB. This leads to paced V_6_ RWPT > native V_6_ RWPT.

The primary aim of this study was to describe electrocardiographic and electrophysiologic characteristics of RBBP in order to establish diagnostic criteria for RBBP.

Secondary aims were to compare pacing parameters and basic echocardiographic and clinical outcomes between RBBP and HBP.

## Methods

The data that support the findings of this study are available from the corresponding author upon reasonable request. The study adhered to the Helsinki Declaration and was approved by the Institutional Bioethical Committee.

### Population

All consecutive patients who underwent CSP device implantation for bradycardia and/or heart failure indications in a single university-based center in the period between May 2014 and January 2022 were screened using a prospectively-maintained CSP database. The following inclusion criteria were applied

1. Pacing lead positioned in the right ventricle / para-annular HB area,
2. Unquestionable conduction system capture confirmed with QRS morphology transition during threshold test or programmed stimulation.^9, 10^
3. His bundle-ventricle (H-V) / right bundle branch-ventricle (RB-V) interval < 55 ms.
4. Baseline conducted QRS other than LBBB.

### Analyzed variables, definitions and measurements

The following hypothetical criteria for identifying RBBP were assessed in different conduction system potential to QRS interval categories:

### 1. Double QRS morphology transition

Since during proximal RBBP, not two but three excitable structures are adjacent (HB, RBB and the septal myocardium), not just a single (as with HBP) but also a double QRS transition may be possible:

- non-selective(ns)-HBP → ns-RBBP → RVSP → loss of capture
- ns-HBP → ns-RBBP → selective(s)-RBBP → loss of capture
- ns-HBP → s-HBP → s-RBBP → loss of capture

QRS transition was defined as change in QRS morphology in several leads accompanied by either the appearance/prolongation of a latency interval, or in a sudden prolongation of V_6_ R-wave peak time (V_6_ RWPT). Examples of different types of double QRS transition are presented in **Figures 3 and 4** and **Supplemental Figures 2-4**.

**Figure 3.**
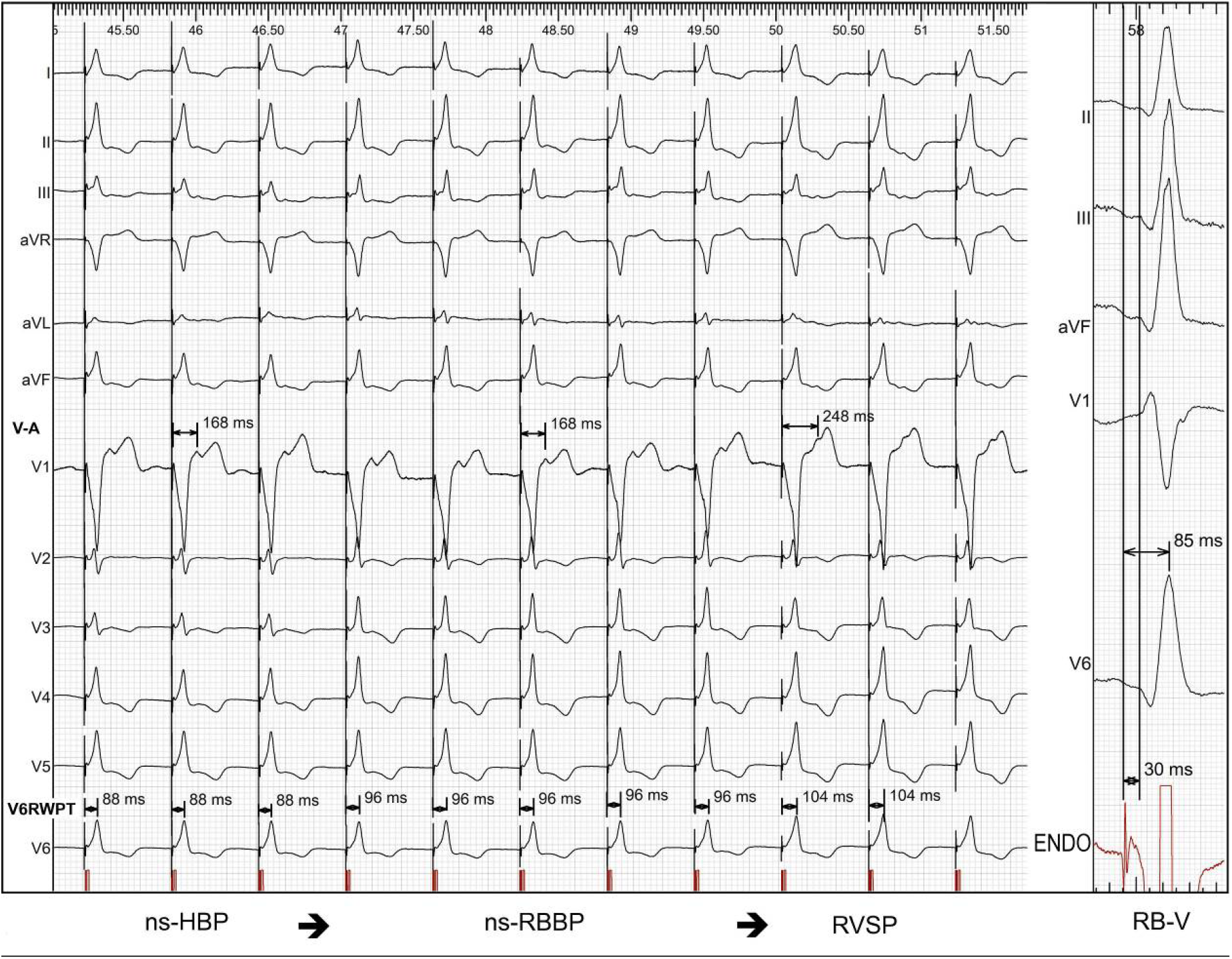
Double QRS transitions during threshold testing. First, there is transition from non-selective His bundle pacing (ns-HBP) to non-selective right bundle branch pacing (ns-RBBP) and then to right ventricular septal pacing (RVSP), as indicated by ventricular-atrial interval prolongation. Each transition results in incremental prolongation of V_6_ R-wave peak time (V_6_ RWPT). Right bundle branch to ventricle (RB-V) interval of 30 ms was recorded at the pacing lead implantation site with potential to V6 R-wave peak of 85 ms. As a result of double QRS transition, there is no single substantial (i.e. > 20 ms) increase of V_6_ RWPT.

**Figure 4.**
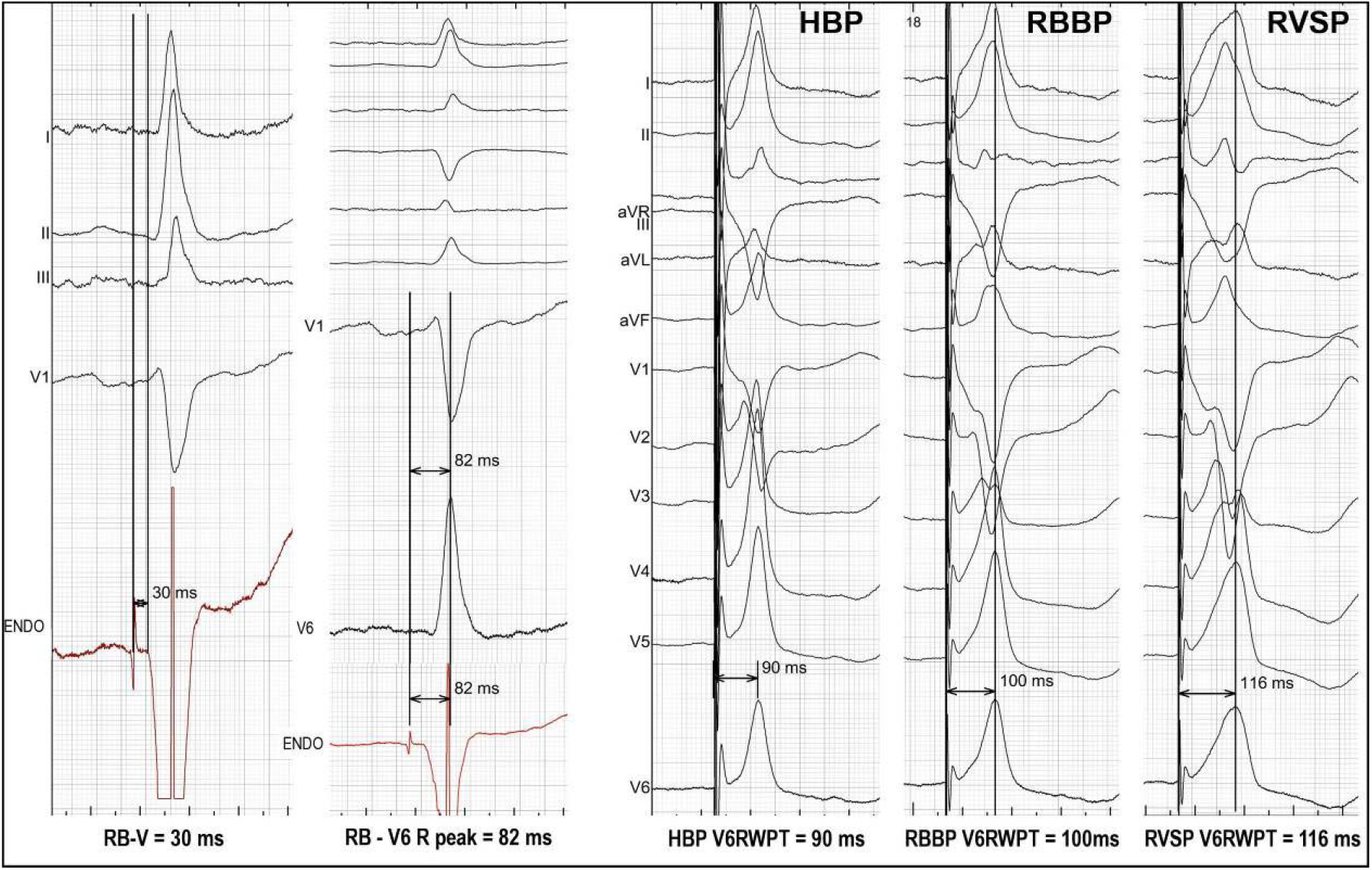
Effect of different types of capture on V^6^ RWPT. Patient with right bundle branch pacing (RBBP), right bundle branch to ventricle interval (RB-V) of 30 ms and potential to V_6_ R-wave peak of 82 ms. Two QRS transitions with small (< 20 ms) increments of V_6_ R-wave peak time (V_6_ RWPT) were present. At high output, His bundle pacing (HBP) was obtained with paced V_6_ RWPT of 90 ms, similar to native potential to V_6_ RWPT of 82 ms (±10 ms). At working output, RBBP was observed, resulting in V_6_ RWPT increase by 10 ms. At low output, loss of RBB capture was observed resulting in right ventricular septal pacing (RVSP) and further increase of V_6_ RWPT by 16 ms.

#### 2. Selective RBBP QRS is different compared to native conducted QRS

In contrast to HBP, during s-RBBP, paced QRS morphology should display features suggestive of delay in LV activation due to the preexcitation of the RBB. Difference in QRS morphology was defined as a visually obvious change in amplitude and/or duration in the precordial leads V_1_ -V_6_. Five illustrative examples are presented in **Figure 5** and **Supplemental Figures 2 and 4**.

**Figure 5.**
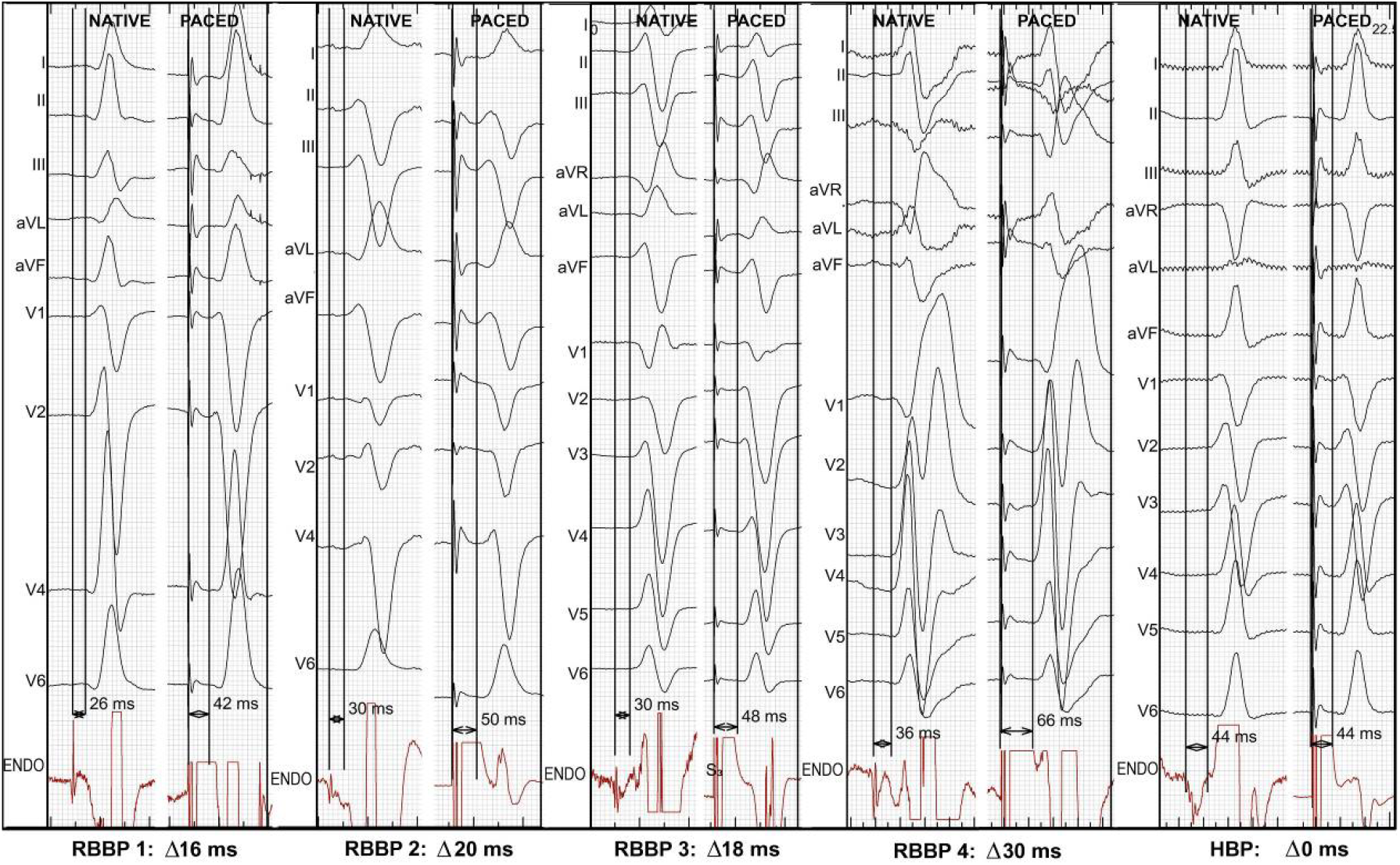
Latency and QRS morphology with selective right bundle branch pacing (RBBP) compared to native conduction in four patients. During selective RBBP, the stimulus to QRS (paced latency)) was longer than the native latency interval (RBB potential to QRS). Difference (Δ) between paced and native latency intervals ranged from 16 to 30 ms, being most pronounced in the patient with right bundle branch block. In the first three patients, the paced QRS complex was different compared to native rhythm; in leads V1-V3, r waves became smaller and S waves deeper. None of these were present during His bundle pacing (HBP).

#### 3. Stimulus-V_6_ RWPT > potential-V_6_ RWPT

In contrast to HBP, during RBBP, the interval from stimulus to V_6_ R-wave peak should be longer than the corresponding native interval measured from the conduction system potential to V_6_ R-wave peak. A difference between paced and native V_6_ RWPT ≥ 10 ms was considered diagnostic. The mechanism of this phenomenon is explained in **Figure 2**.

#### 4. Stimulus-sQRS > potential-QRS

The mechanism of this phenomenon of latency interval during selective pacing being longer than RB-V interval is explained in **Supplemental Figure 1**.

#### 5. Increase in V_6_ RWPT with loss of HB/RBB capture is small

An increase in V_6_ RWPT during transition from ns-RBBP to RVSP is small (< 20 ms) because of baseline delay in LV activation during RBBP. The same is expected with transition from ns-HBP to ns-RBBP, due to maintained conduction system activation.

#### 6. Capture thresholds of the conduction system and adjacent septal myocardium are equal

In contrast to HBP, more distal conduction system pacing (e.g. LBBP) typically results in obligatory simultaneous activation of the adjacent myocardium. Equal capture threshold during threshold test was defined as a direct transition from non-selective QRS to loss of capture without the phase of selective CSP or selective myocardial capture (one such beat before loss of capture was allowed).

All procedures were recorded on a digital electrophysiological system (LabsystemPRO, Boston Scientific, USA). Conduction system potential was recorded by the pacing lead connected to the electrophysiological system. To ensure high precision, the measurements were performed using all 12 surface ECG leads and the endocardial channel recorded simultaneously, digital calipers and fast sweep speed (200 mm/s).

In every patient, each observed paced QRS type (s-HBP, ns-HBP, s-RBBP, ns-RBBP and RVSP) as well as native conducted QRS were analyzed. Global QRS duration was measured(interval from the earliest onset to the latest offset of the QRS in all 12 leads recorded simultaneously; for paced QRS, pacing stimulus was considered as QRS onset). To ensure precision, at least ten intervals were measured for each patient and their values averaged.

The working pacing output was defined as the pacing output in the range of 3.0 – 3.6 V at 0.4 ms pulse width.

## Results

### Population

A total of 1412 patients who underwent conduction system pacing device implantation were screened and 403 HBP/RBBP patients were identified. Out of this group, 326 patients fulfilled the inclusion criteria and were further analyzed, while the remaining 77 were excluded mainly due to the baseline HV interval > 55 ms or LBBB (**Supplemental Figure 5)**.

Baseline clinical characteristics of the studied cohort are presented in **Table 1**.

**Table 1.** Basic characteristics of the studied group

|  | Potential-QRS < 35 ms<br>n = 53 | Potential-QRS = 35-55 ms<br>n = 273 | p-value |
| --- | --- | --- | --- |
| Age [years] | 75.3 ± 12.6 | 75.1 ± 11.1 | 0.89 |
| Females | 19 (35.8%) | 113 (41.4) | 0.45 |
| Pacing indication [n]: |  |  |  |
| • Sick sinus syndrome | 12 (22.7%) | 77 (28.2%) | 0.74 |
| • Atrioventricular block | 14 (26.4%) | 63 (23.1%) |  |
| • Atrial fibrillation | 21 (39.6%) | 90 (33%) |  |
| • Heart failure | 6 (11.3%) | 43 (15.7%) |  |
| Comorbidities [n]: |  |  |  |
| • Diabetes mellitus | 15 (28.3%) | 90 (33%) | 0.6 |
| • Coronary heart disease | 17 (32.1%) | 91 (33.3%) | 0.86 |
| • Heart failure | 16 (30.2%) | 118 (43.2%) | 0.08 |
| • Hypertension | 41 (77.4%) | 231 (84.6%) | 0.19 |
| • Severe valvular disease | 2 (3.8%) | 22 (8.1%) | 0.27 |
| Native QRS duration [ms] | 108.6 ± 22.6 | 111.5 ± 20.8 | 0.36 |
| Native QRS type: |  |  |  |
| • Narrow | 31 (58.5%) | 157 (57.5%) | 0.56 |
| • LAFB | 4 (7.5%) | 13 (4.8 %) |  |
| • Incomplete RBBB | 3 (5.7%) | 8 (2.9%) |  |
| • RBBB | 4 (7.5%) | 29 (10.6%) |  |
| • RBBB + LAFB/LPFB | 5 (9.5%) | 12 (4.4%) |  |
| • NIVCD | 6 (11.3%) | 54 (19.8%) |  |

RBBB – right bundle branch block; LAFB – left anterior fascicular block; LPFB – left
posterior fascicular block; NIVCD – non-specific intraventricular conduction disturbance.

### RBBP prevalence and criteria

In the whole studied cohort of 326 patients, RBBP -as per the diagnostic criteria described below -was observed in 64 (19.6%) patients. RBBP only was observed in 36 (11.0%) patients, both RBBP and HBP in 28 (8.6%) and HBP only in 262 (80.4%). In patients with potential to QRS in the HB range (35-55 ms), RBBP was observed mainly at near-threshold values, while in patients with potential to QRS interval in the RBB range (< 35 ms), RBBP was usually observed at working output (**Table 2**).

**Table 2.** Right bundle branch pacing (RBBP) criteria and capture and working output in different conduction system potential to QRS interval categories.

|  | **P-V**<br>**≤ 30 ms**<br>**n = 31** | **P-V**<br>**= 31-34 ms**<br>**n = 22** | **P-V**<br>**= 35-39 ms**<br>**n = 73** | **P-V**<br>**= 40-55 ms**<br>**n = 200** | **p-value** |
| --- | --- | --- | --- | --- | --- |
| Double QRS transition | 7 (22.6%) | 8 (36.4%) | 6 (8.2%) | 7 (3.5%) | < 0.001 |
| Selective paced QRS ≠ native QRS | 7 (22.6%) | 11 (50%) | 9 (12.3%) | 15 (7.5%) | < 0.001 |
| Stimulus-V<sub>6</sub>RWPT > potential-V<sub>6</sub>RWPT | 29 (93.5%) | 19 (86.4%) | 8 (11%) | 8 (4%) | < 0.001 |
| Stimulus-sQRS > potential-QRS | 14 (45.2%) | 11 (50%) | 7 (9.6%) | 11 (5.5%) | < 0.001 |
| Delta V<sub>6</sub>RWPT ≤ 20 ms | 14 (45.2%) | 9 (40.9%) | 3 (4.1%) | 3 (1.5%) | < 0.001 |
| Equal capture thresholds | 12 (38.7%) | 5 (22.7%) | 4 (5.8%) | 10 (5%) | < 0.001 |
| RBBP at working output | 21 (67.7%) | 14 (63.6%) | 6 (8.2%) | 1 (0.5%) | < 0.001 |

P-V – His bundle/right bundle branch potential to QRS interval
V<sub>6</sub>RWPT – V<sub>6</sub> R-wave peak time.

Double QRS transition (which initially served as the gold standard for RBBP diagnosis) was observed in all potential to QRS interval categories, although much more commonly with shorter intervals (**Table 2**). In patients with potential to QRS in the HB range (33-55 ms) double transition was predominantly observed at low output. Consequently, RBBP at working output was very rare. In contrast, in patients with potential to QRS in the RBB range, RBBP was often present at working output (**Table 2**).

When there was QRS transition from HBP to RBBP (n = 28), the criterion of stimulus-V_6_ RWPT > potential-V_6_ RWPT (Δ ≥ 10 ms) always discriminated between HBP and RBBP (100% sensitivity). Apart from these 28 cases, this criterion identified an additional 36 RBBP cases without HBP present even at high amplitude (i.e. without double QRS transition). Almost all of these cases (93.3%, 33/36) had potential to QRS in the RBB range (< 35 ms) – **Table 2**. Furthermore, in all of these cases, at least one more RBBP criterion was present with a median of two criteria per patient. In patients without any other RBBP criteria and definite HBP, the criterion was never fulfilled (100% specificity). Consequently, the criterion of stimulus-V_6_ RWPT > potential-V_6_ RWPT was accepted as the new diagnostic standard for RBBP and further categorization of patients and assessment of other parameters and criteria were based on this criterion. The average difference between of paced V_6_ RWPT and native V_6_ RWPT in RBBP cases was 17.8 ms (98.2 ±8.8 ms vs 80.3 ±8.4 ms, p < 0.001) while the difference in HBP cases was only 0.5 ms (92.6 ±10.7 ms vs 92.1 ±10.5 ms, p = 0.07). ROC curve analysis confirmed that the arbitrary cut-off for ΔV_6_ RWPT ≥ 10 ms was optimal (**Supplemental Figure 6**).

In cases where selective RBB capture was observed, stimulus to QRS (paced latency) was much longer than potential to QRS (native latency): 52.1 ±8.8 ms vs 34.1 ±6.5 ms, p <0.0001, respectively. In contrast, during selective HBP, native and paced latency intervals closely corresponded, with an average difference of 4.1 ms: 43.7 ±5.5ms vs 47.8 ±6.5 ms, p <0.0001, respectively. The criterion of Δ latency ≥ 10 ms was fulfilled in 87.2% (34/39) of RBBP cases. ROC curve analysis confirmed that Δ latency ≥ 10 ms is optimal to differentiate between HBP and RBBP with AUC, sensitivity and specificity of 94.4%, 87.2% and 95.3%, respectively (**Supplemental Figure 6**).

Transition from ns-RBBP to RVSP resulted in less pronounced prolongation of V_6_ RWPT than transition from ns-HBP to RVSP, increase by 21.4 ms (97.7 ±7.6 to 119.2 ms ±11.8, p < 0.0001) vs increase by 34.7 ms (93.6 ±10.1 ms to 128.4 ±17.3 ms, p < 0.0001) respectively. The criterion of V_6_ RWPT increase ≤ 20 ms during QRS transition (considering both ns-HBP to ns-RBBP and ns-RBBP to RVSP) was characterized by area under the ROC curve, sensitivity and specificity for diagnosis of RBBP of 0.888, 70.5% and 94.8% (**Supplemental Figure 6**).

Equal capture thresholds of myocardium and conduction system were more common in patients with RBBP than in patients with HBP: 31.2% (20/64) vs 4.2% (11/262), p < 0.001, respectively. Sensitivity and specificity of this criterion for diagnosis of RBBP was 31.3% and 95.8%, respectively.

### ECG during RBBP

Selective paced QRS morphology differed from native conducted QRS in 64.1% (25/39) s-RBBP cases; these were all sRBBP cases without baseline RBBB (25/25). During RBBP, features of LV activation delay appeared in precordial leads: r wave reduction and S wave increase in leads V_1_ -V_3_, delayed V_6_ R-wave peak and global QRS prolongation. In contrast, s-HBP QRS was different from native conducted QRS only in 8.9% (17/191) cases. Consequently, the criterion of selective paced QRS differing from native conducted QRS had sensitivity and specificity for RBBP of 64.1% and 91.1% respectively. In all cases of selective RBBP it was observed only at low / near threshold output. Consequently, at working output, RBBP was always non-selective.

Transition from ns-HBP to ns-RBBP resulted in V_6_ RWPT prolongation by 11.6 ±4.3 ms, (86.6 ±6.9 ms vs 98.2 ±9.4 ms, p < 0.0001) and QRS prolongation by 10.5 ±5.4 ms, (133.2 ±13.3 ms vs 143.7 ±13.6 ms, p < 0.0001).

Procedural and clinical outcomes results are shown in **Table 3**.

**Table 3.**
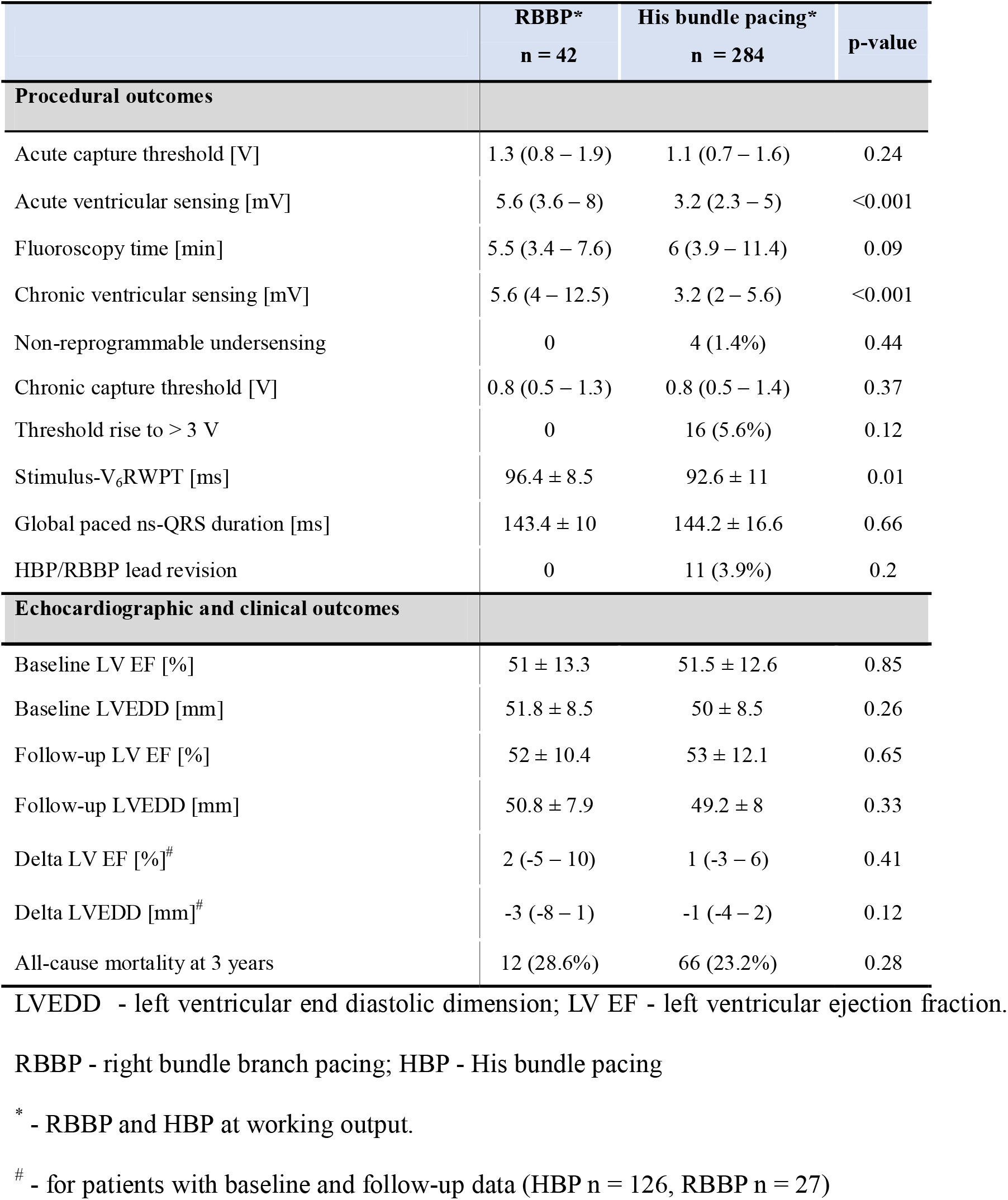
Right bundle branch pacing (RBBP) electrical parameters and outcomes

## Discussion

In this series of patients in whom HBP had been targeted, a distal position of the pacing lead as indicated by a short (<35 ms) potential to QRS interval was relatively frequently observed -in 16.2% (53/326) cases. Evidence of RBBP at any output was observed in 19.6% of patients, while at working output, RBBP was present in 12.2% patients.

The major findings of the study are:

1. RBBP is characterized by distinct electrophysiological and ECG features that differentiate it from HBP.
2. RBBP offers favorable pacing characteristics: good sensing, capture thresholds which are comparable to HBP, and myocardial ‘back up’ capture at working output in all patients.

### Physiology behind the criteria for RBBP

#### Stimulus-V_*6*_ *RWPT > potential-V*_*6*_ RWPT

This electrophysiological criterion, seems to be the best discriminator between HBP and RBBP. A difference of ≥ 10 ms in the V_6_ RWPT intervals was found to have 100% diagnostic accuracy. With HBP, the depolarization pathway from HB to the left ventricle is the same during pacing as during native conduction. Consequently, the paced and native V_6_ RWPT during HBP are equal.^8^ The same rule is valid for LBBP.^7^ In contrast, during RBBP the depolarization wavefront has to travel retrogradely from RBB to HB, then traverse to LBB predestined fibers within the HB, and then to travel anterogradely to reach the LV myocardium (**Figure 3**). Therefore, the difference between paced and native V_6_ RWPT intervals is more pronounced than the few milliseconds of difference between H-V and RB-V intervals. This novel physiologic criterion can be used to differentiate HBP from RBBP during permanent pacing and also during electrophysiologic study.

#### Stimulus to s-QRS > potential-QRS

This criterion is illustrated by **Figure 5** and **Supplemental Figure 1**. During selective HBP the interval from stimulus to QRS onset (paced latency) is nearly identical to the H-V interval (native latency), whilst during RBBP, it is always longer than RB-V interval. In patients with RBBB distal to the pacing lead, explanation of this phenomenon follows the same simple logic as for V_6_ RWPT, that ventricular activation depends then purely on retrograde conduction from RBB to LBB. Counterintuitively, in patients with preserved RBB conduction, the RB-V interval during RBBP is also determined by LBB conduction, because it is LBB activation of the interventricular septum which is responsible for QRS onset. This is because there is a difference between LBB and RBB conduction times. This difference is in the range of 5 – 20 ms, as indicated by H-V interval prolongation during acute development of LBBB.^11-13^ It is this difference that causes and determines the upper limit of the latency interval prolongation observed during RBBP.

#### Potential to QRS interval

The commonly accepted lower range of normal value for HV interval of 35 ms seems to be a good initial discriminator between HBP and RBBP. However, a fixed cut-off of 35 ms does not take into account the variability in HB length, RBB take-off level and decreased velocity of conduction in the diseased conduction system. Also, more importantly, the H-V / RB-V interval recorded from the pacing electrode only localizes lead position along the conduction system. However, it does not inform which structure is captured. Electrophysiological and ECG criteria for RBBP might be more informative than fixed cut-offs for potential to QRS interval. For example, out of 53 patients with potential to QRS interval < 35 ms, HB at any output was present in 5 (9.4%), RBBP only at near threshold output in 13 (24.5%) and RBBP at working output in 35 (66.0%) -Table 1. Most likely, in cases with HBP, despite recording of RBB potential, the pacing lead is positioned in the area of the most proximal RBB and the size of the virtual electrode at working output is big enough to result in simultaneous direct capture of the HB, or the RBB takeoff is more distal than usual. In 2.6% (7/273) patients, the inverse situation was observed: despite potential to QRS interval ≥ 35 ms there was RBB capture at working output. This could be explained by an earlier take-off of the RBB or slower conduction in the RBB (the presumed HB potential was in fact an RBB potential) or longitudinal HB dissociation with a higher capture threshold of the HB fibers predestined to LBB. In an additional 3.3% (9/273) patients, RBBP was observed only at low output, probably indicating preferential activation of RBB fibers within HB with low energy/small virtual electrode.

### ECG during RBB capture

Ample myocardium around the RBB always leads to non-selective capture at working output. Preexcitation of the RBB results in relative delay in LV activation in comparison to native conduction. This manifests itself by a wider QRS complex and longer V_6_ RWPT during RBBP. Consequently, s-RBBP QRS is different than native conducted QRS, while nonselective RBBP QRS is intermediate between HBP and RVSP (**Figures 3 and 4**) with a pronounced pseudo-delta wave. In patients with complete RBBB, the QRS during s-RBBP and HBP are identical; however, latency during selective RBB capture might be exceptionally long (**Figure 5**).

During transition from ns-HBP to RVSP the V_6_ RWPT increases by the time it takes to cross the interventricular septum and engage the LBB, this is on average 35 ms and nearly always > 20 ms.^8^ During RBBP, the V_6_ RWPT is already long and any additional prolongation with transition to RVSP is, therefore, shorter. Small difference in paced V_6_ RWPT between ns-RBBP and RVSP can make it challenging to differentiate between these two capture types.^14^

#### Clinical translation

Proximal RBBP can potentially address some of the drawbacks of HBP and LBBP. Ample amount of myocardium around the RBB ensures good sensing and “backup” myocardial capture thresholds. RBBP is similar to LBBP in this respect. Interestingly, no lead related problems (undersensing, oversensing, late threshold rise) were observed in patients with RBBP at working output. Furthermore, in case of loss of RBB activation, the para-RBB lead position is more midseptal with probably faster access to the distal conduction system. RBBP does not require invasive ventricular transseptal access to the conduction system (in contrast to LBBAP). This may facilitate lead extraction and may also be pertinent for future leadless pacing that targets the conduction system.

Disadvantages of RBBP are related to more delayed left ventricular activation in comparison to HBP and LBBP. Nevertheless, the RBBP QRS is narrower and more physiological than with RVSP. This might be enough to prevent pacing-induced cardiomyopathy, as RBBP paced QRS duration is usually < 150 ms, but is likely to be insufficient to obtain optimal hemodynamic and clinical response in heart failure patients. Moreover, RBBP is unable to correct the conduction defect in LBBB patients, making it unsuitable as the sole pacing mode for most cardiac resynchronization candidates. However, it might be an excellent pacing modality when combined with LBBP as a part of dual branch pacing system to fully restore physiological depolarization.^15^ Similarly, combing RBBP with coronary venous pacing could make it possible to optimally synchronize left and right ventricular activation in patients in whom fusion pacing with intrinsic RBB conduction is not feasible (e.g. in case of atrial fibrillation).

Long-term echocardiographical outcomes and all-cause mortality between RBBP and HBP in our study seem comparable (also see **Supplementary Material**). However, prospective studies assessing clinical outcomes of RBBP in greater detail are warranted.

### Limitations

This is single center, retrospective observational study with all inherent limitations of such design. However, large studied sample, prospectively maintained database, high technical standard of ECG and electrophysiological data acquisition and measurements ensures reliability of the reported results.

## Conclusions

This study provides the first description and evaluation of permanent RBBP. Novel electrophysiologic and ECG-based diagnostic criteria for this distinct conduction system pacing modality were formulated and assessed. The current study adds mechanistic insights to our understanding of the ECG and physiology of CSP.

RBBP is probably an underrecognized and underutilized pacing modality that might be optimal in some CSP candidates.

## Data Availability

All data produced in the present study are available upon reasonable request to the authors

