## Supplementary Material for "Right bundle branch pacing: criteria, characteristics and outcomes"

**Supplementary methods**

Echocardiography examination.

An echocardiographic examination was performed at baseline and later periodically at follow-up visits. Examinations were conducted using standard ultrasound system (Vivid 7 and Vivid E9, General Electrics, Norway). Four consecutive cardiac cycles with constant heart rate were recorded and all images were stored digitally for later off-line analysis using EchoPac software (v293, General Electrics, Norway). According to the current recommendations, the 2D echocardiography assessment included parasternal long- and short-axis views, and apical 4-, 3- and 2-chamber views. Left ventricule ejection fraction was calculated based on Simpson’s biplane method.

Mortality status

After follow-up appointment for which the patient failed to come, the patient/patient’s family was contacted to check mortality status.

Statistical analysis

Continuous variables are presented as means and standard deviations. Categorical variables are presented as percentages. Between-group differences were assessed using contingency tables, Student’s t-test, or analysis of variance, as appropriate. The performance of binary decision rules was described using sensitivity and specificity. The performance RBBP criteria and optimal cut-offs to differentiate between HBP and RBBP was assessed using the receiver operating characteristic (ROC) curve. Statistical analysis was performed using SPSS statistical software (IBM Statistics 27; Chicago, IL, USA).

**Supplementary Results**

ECG during RBBP

The QRS during RVSP from the para-RBB region with loss of RBB capture was more narrow than with para-HB RVSP, 165 ±14.6 ms and 176 ±21.5 ms, p = 0.004, respectively. Moreover, RVSP from the para-RBB region was characterized by faster LV activation as indicated by shorter V_6_RWPT than with para-HB RVSP: 119.1 ±11.8 ms and 128 ±17.3 ms, p = 0.004, respectively

Long-term echocardiographic outcomes and all-cause mortality.

Echocardiographic data were available for 93.6% patients at baseline and for 48.2% patients during follow-up. No significant differences between RBBP and HBP (categorized per capture type at working output) outcomes were found (**Table 3**).

At the 3-year follow-up 78 patients had died. There was no relation between mortality and the RB-V/H-V interval and no relation to the pacing type at the working output (**Table 3**): all-cause 3-year mortality rate in RBBP and HBP of 28.6% and 23.2% (p = 0.28) with RBBP and HBP, respectively.

**Supplementary Discussion**

With myocardial-only capture, the more mid-septal RBBP lead position compared to with HBP results in shorter V_6_RWPT, adding to reduced difference in V_6_RWPT during transition from non-selective capture to myocardial-only capture.

RBBP in patients with baseline incomplete RBBB results in more balanced RV and LV activation and attenuation of RBBB features – this may be mistaken with direct RBBB correction by the pacing stimulus (**Supplementary Figure 7**).

**Supplementary Figures**

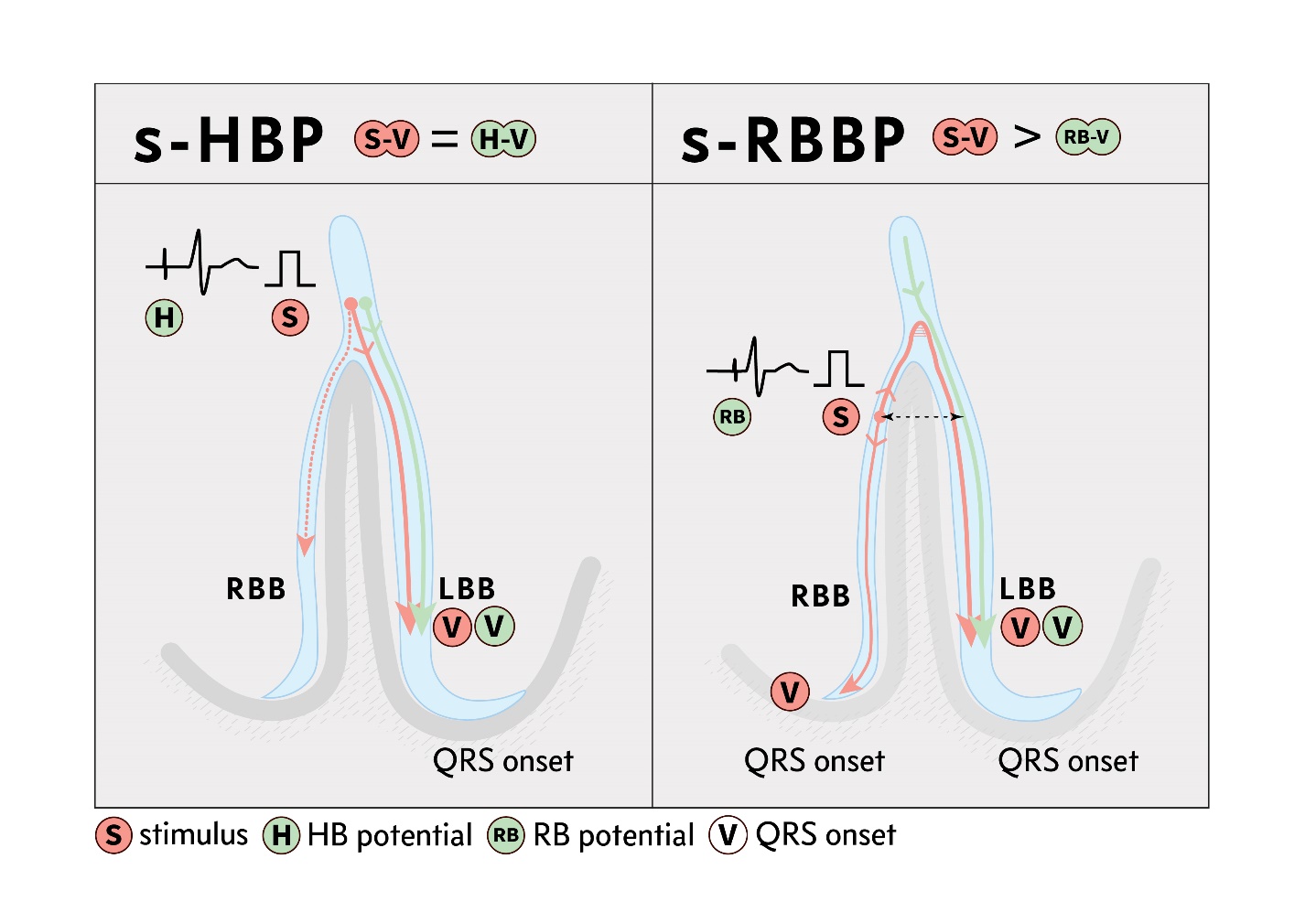

**Supplemental Figure 1**. Difference between latency intervals with pacing and native conduction during selective His bundle pacing (s-HBP) and selective right bundle branch pacing (s-RBBP).

Left panel: during s-HBP (red line), the activation pathway from the point of HB capture (S) or HB potential recording (H) to the earliest activation of the myocardium (V) occurring via the LBB is the same during pacing and intrinsic conduction (green line), leading to S-V = H-V.

Right panel: during selective RBBP, activation of the myocardium can occur either via retrograde conduction to the LBB or via anterograde conduction in the RBB. Regardless of which of these two routes is faster and is responsible for QRS onset, the interval from the stimulus to QRS onset (S-V) will be longer than RBB potential to QRS onset (RB-V). This is counterintuitive because the RB-V interval is, in way, artificially short, as it does not reflect the conduction from RBB to the right ventricle for the reason that QRS onset results from the simultaneous faster activation of the septum via LBB. This is because there is a difference between LBB and RBB conduction times of approximately 5-20 ms. Therefore, during selective RBBP, the RB-V interval is not reproduced, but the paced latency is longer reflecting retrograde conduction to the LBB and/or long anterograde conduction in the RBB to the right ventricle.

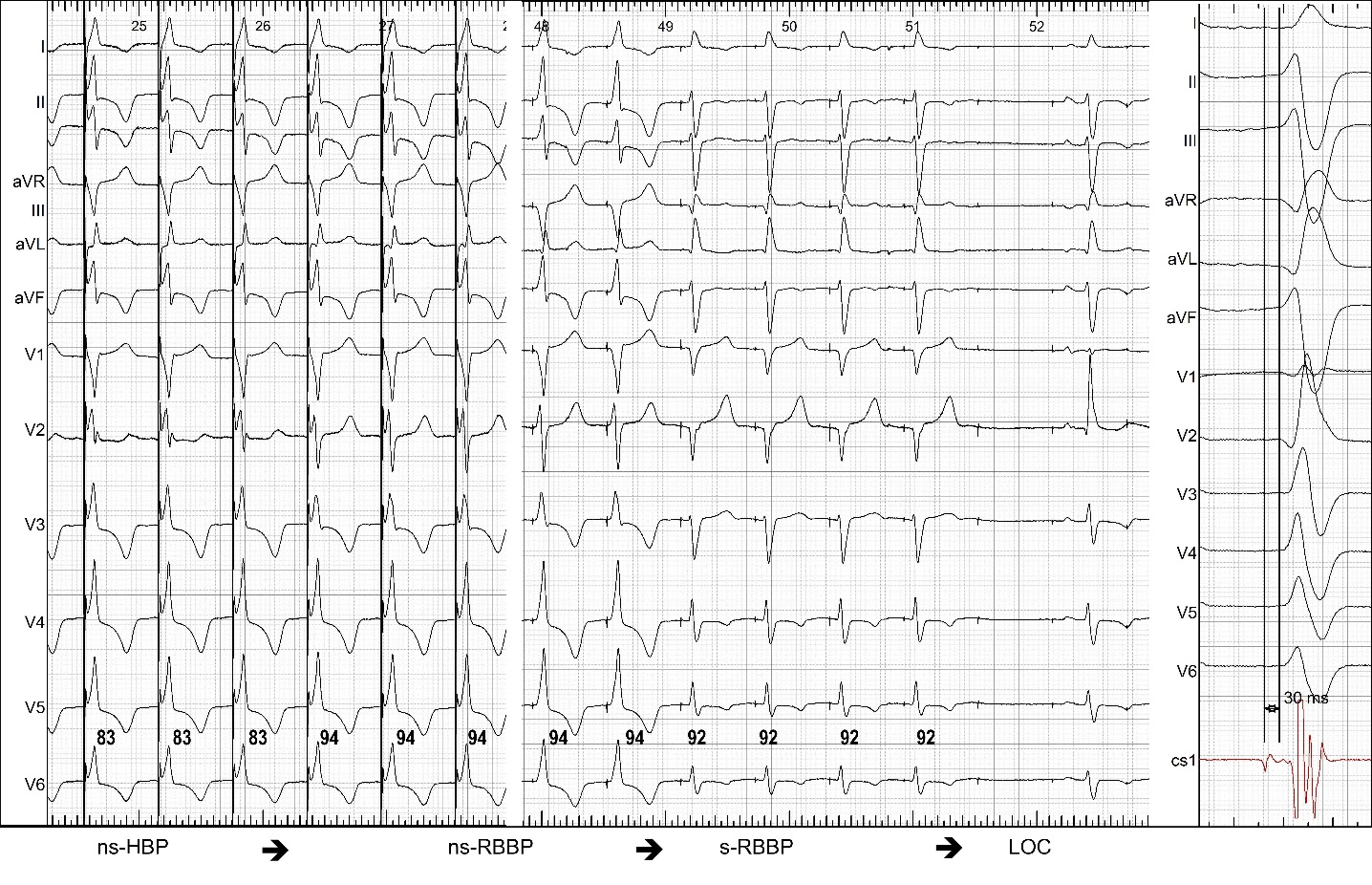

**Supplemental Figure 2**. Double QRS transition during threshold test with unipolar pacing. Non-selective His bundle pacing (ns-HBP) transitions to non-selective right bundle branch pacing (ns-RBBP) with V_6_ R-wave peak time (V_6_RWPT) prolongation of 11 ms (there is also a slight change in overall QRS morphology – see lead V2). Second transition is from ns-RBBP to selective (s-) RBBP. Pseudo-shortening of V_6_RWPT by 2 ms is related to change of R morphology to rS morphology in V_6_. The right bundle branch potential to QRS interval of 30 ms recorded at the pacing lead implantation site is much shorter than latency interval during s-RBBP with a stimulus to QRS interval of 66 ms. Moreover, s-RBBP QRS morphology is different from native QRS (see leads V1-V3) resulting from relative delay in left ventricular activation.

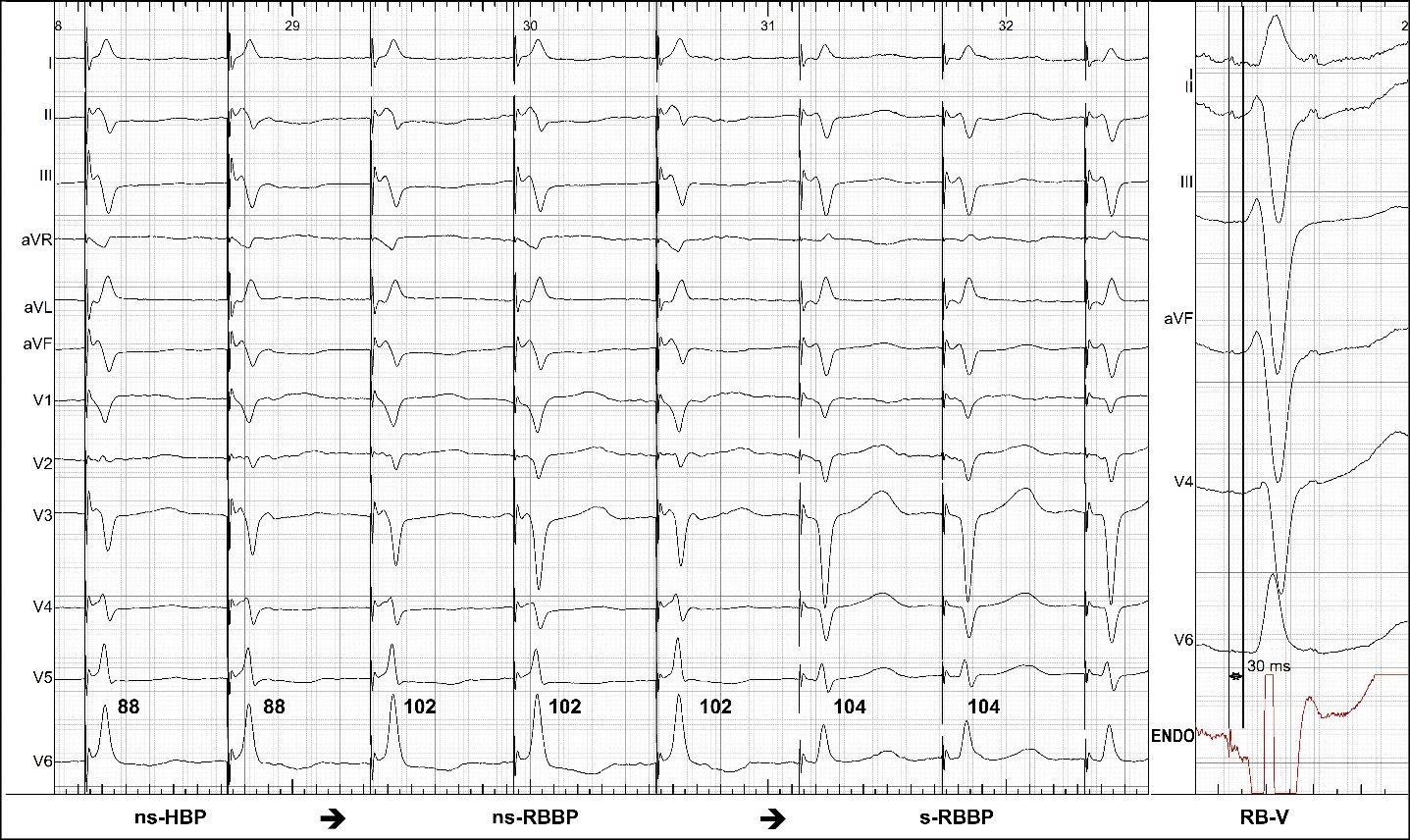

**Supplemental Figure 3**. Double QRS transition during threshold test. Non-selective His bundle pacing (ns-HBP) transitions to non-selective right bundle branch pacing (ns-RBBP) with V_6_ R-wave peak time (V_6_RWPT) prolongation of 14 ms. Second transition is from ns-RBBP to selective (s-) RBBP. Right bundle to QRS interval of 30 ms was recorded at the pacing lead implantation site while stimulus to QRS interval during s-RBBP is 48 ms (not shown).

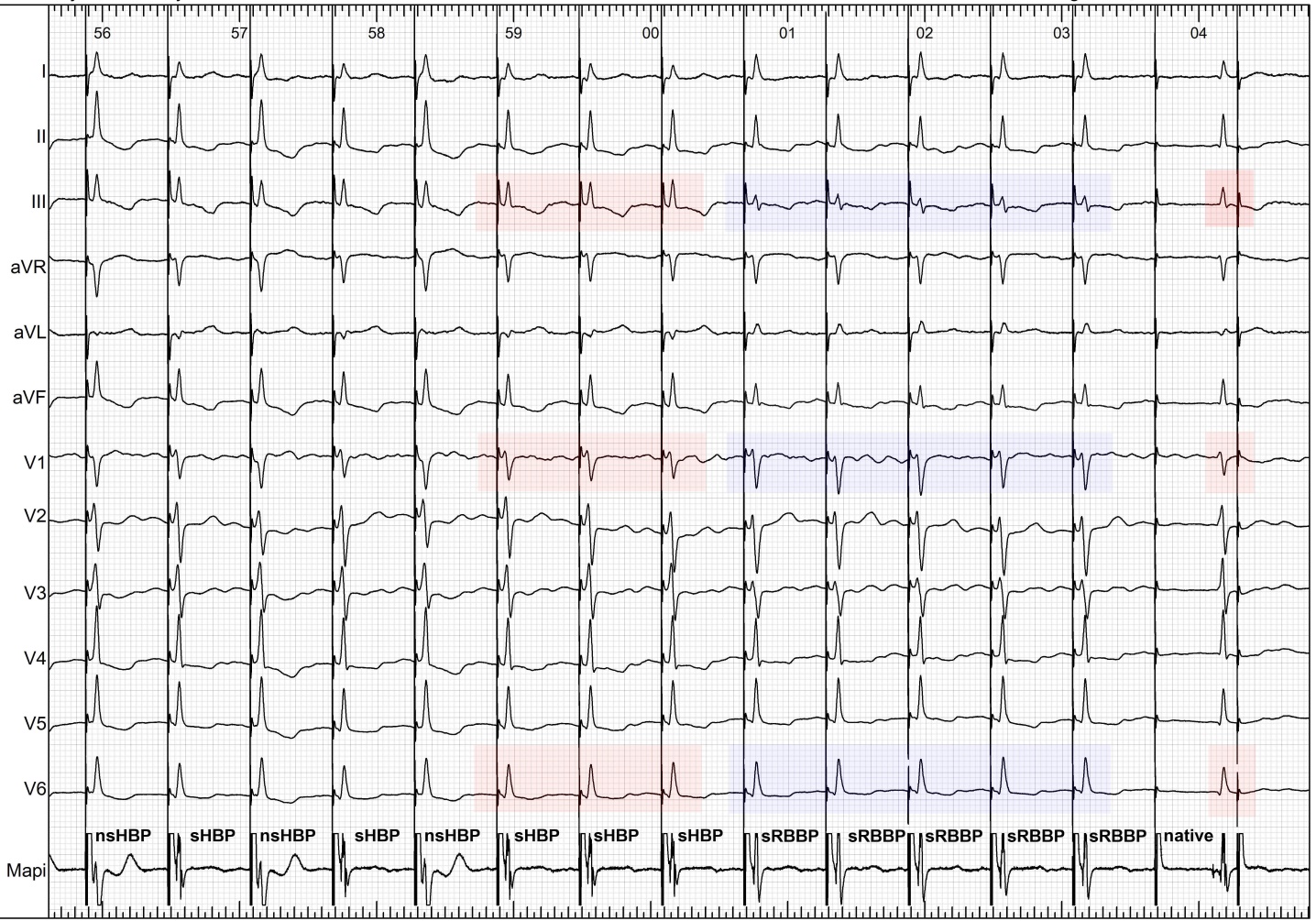

**Supplemental Figure 4**. Double QRS transition during threshold test**.** Non-selective His bundle pacing (ns-HBP) transitions to selective His bundle pacing (s-HBP). Second transition is from s-HBP to s-RBBP. Notably, s-HBP QRS is identical to native rhythm, while s-RBBP QRS has different axis and smaller r waves and deeper S waves in leads V_1_-V_3_, longer V_6_ R-wave peak time (95 ms vs 85 ms, not shown) and latency interval of 45 ms that was longer than the native latency (potential to QRS interval) of 29 ms (not shown).

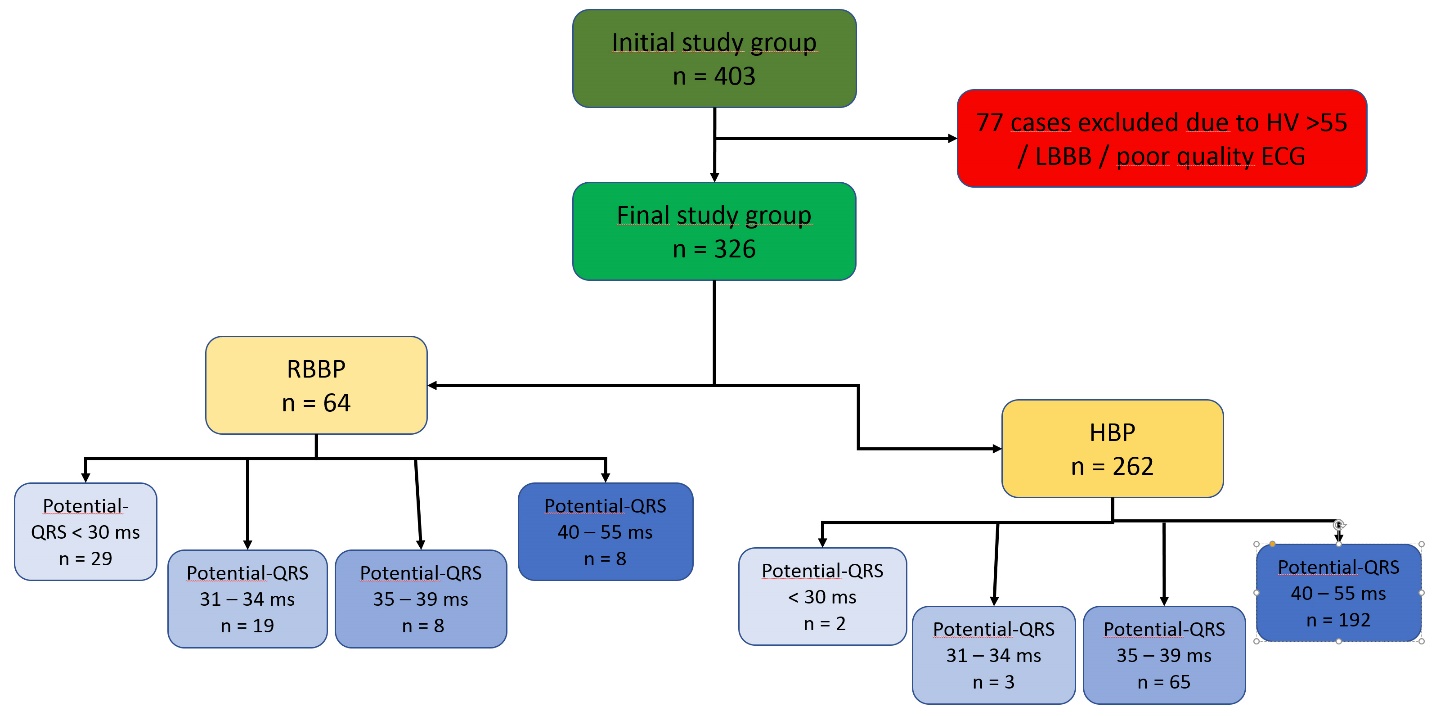

**Supplemental Figure 5**. Study flowchart and capture type in relation to conduction system potential to QRS interval. LBBB – left bundle branch block; RBBP – right bundle branch pacing; HBP – His bundle pacing

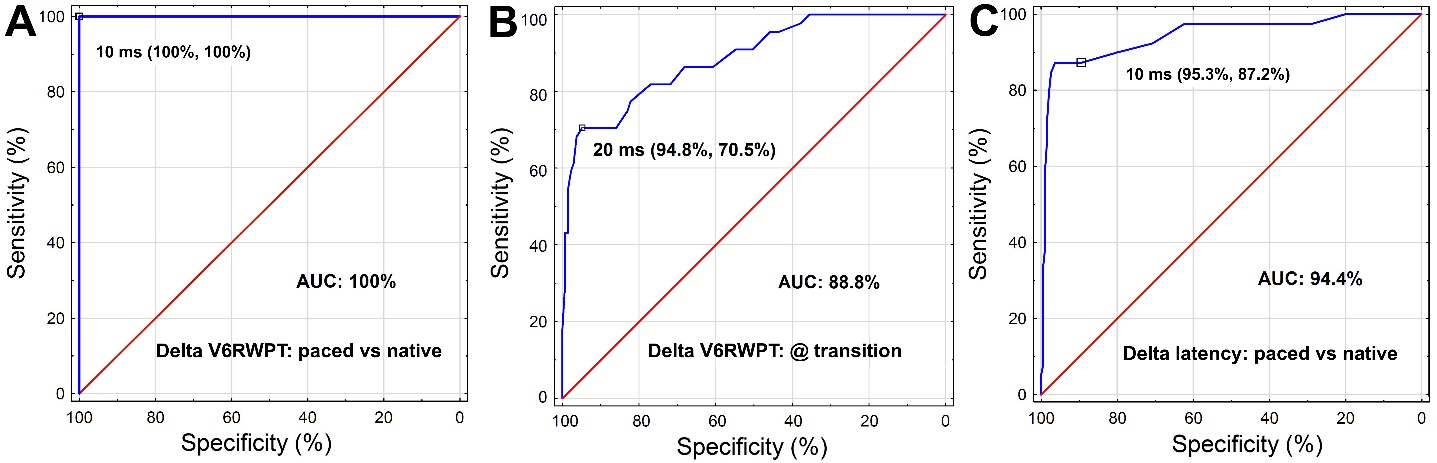

**Supplemental Figure 6**. The receiver operating characteristic (ROC) curve for diagnosis of RBBP using: A) difference ≥ 10 ms between paced V_6_ R-wave peak time (V_6_RWPT) and native V_6_RWPT measured from the stimulus and conduction system potential, respectively, B) V_6_RWPT increase with paced QRS transition during threshold test, C) difference between paced and native latency intervals, measured from the conduction system potential to QRS and stimulus to QRS, respectively.

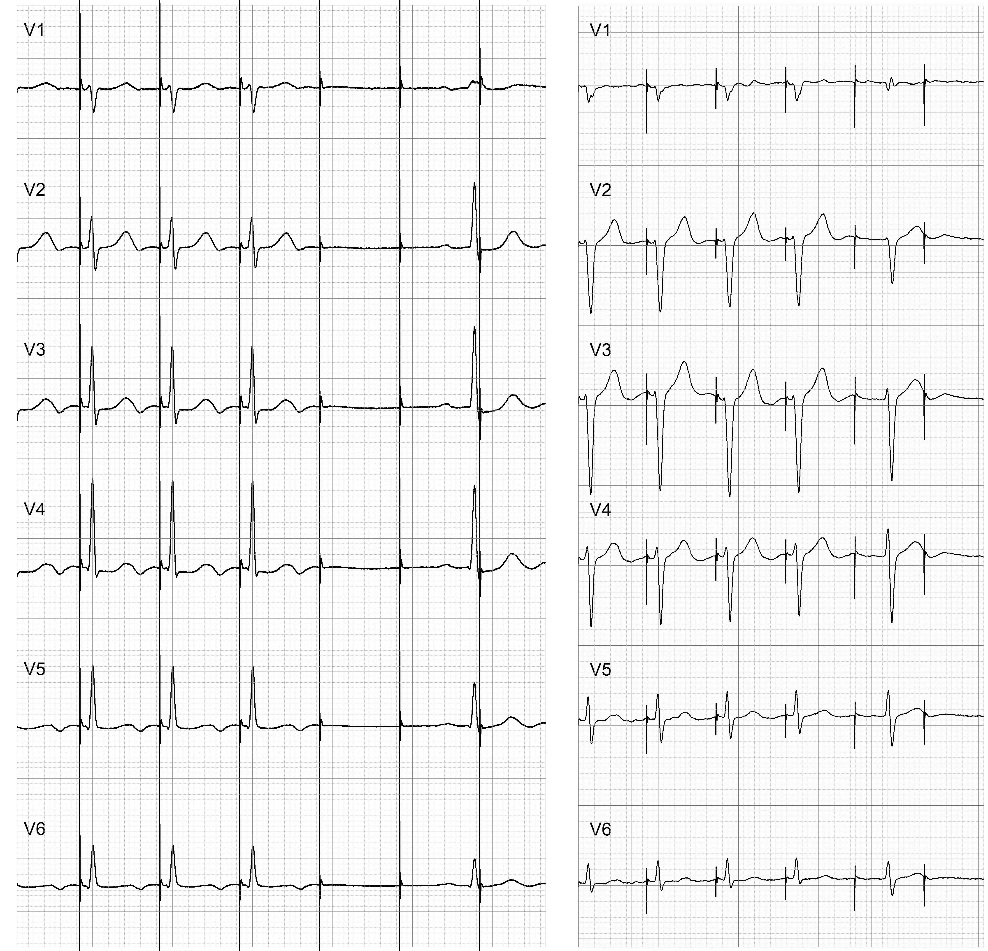

**Supplemental Figure 7**. Two examples of right bundle branch pacing (RBBP) in patients with baseline delay in the right bundle branch (RBB). Preexcitation of the RBB during RBBP results in QRS normalization. This could be mistaken for RBBB correction via recruitment of the diseased RBB fibers. However, correction of RBBB at near threshold outputs (as indicated by loss of capture by subsequent stimuli in these cases) would not be present.
